# Heat and Emergency Department Operations: A Two-Decade Observational Study

**DOI:** 10.64898/2026.10.27.26366595

**Authors:** Neil Singh Bedi, Quinn H. Adams, Bryan Stenson, Natalie Jansen, Gregory A. Wellenius, Caleb Dresser

## Abstract

Emergency department (ED) crowding remains one of the most pressing challenges facing the US healthcare system; environmental factors have received little attention as determinants of ED operational performance, patient behavior, or emergency service utilization. We examined the association between maximum daily heat index and ED throughput metrics, patient behavioral outcomes, and likelihood of EMS arrival. We retrospectively analyzed 485,403 warm-season ED visits from a 22-year period at a large urban academic medical center in New England. The exposure was daily maximum heat index (ERA5-Land) for Suffolk County, Massachusetts. Primary outcomes included four ED throughput time metrics: door-to-doctor time, doctor-to-decision time, boarding time, and total ED length of stay; secondary outcomes included the left without being seen rate, leaving against medical advice rate, and likelihood of EMS arrival. Each 1°F increase in heat index was associated with a 0.41% increase in door-to-doctor time (95% CI: 0.38% – 0.44%). Heat index was not significantly associated with the other three throughput metrics. At the 95°F heat index threshold, patients were 27% more likely to leave against medical advice than on days below the threshold (OR 1.27; 95% CI: 1.02 – 1.58), and had higher odds of arriving by EMS (OR 1.10; 95% CI: 1.06 – 1.15). Higher heat index was associated with longer door-to-doctor time, increased EMS arrival proportion, and higher odds of leaving against medical advice. Health systems and municipalities should characterize how hot weather affects operational performance and integrate this evidence into heat-readiness planning.

## INTRODUCTION

Emergency department (ED) overcrowding represents one of the most pressing healthcare challenges in the United States.^1^ Growth in ED visit volumes over recent decades, combined with insufficient inpatient and post-acute care capacity and throughput, has outpaced available resources, resulting in longer wait times, strained staffing, and widening gaps in the capacity to deliver timely emergency care.^1,2^ The clinical consequences of ED crowding are well-established, with crowding independently associated with prolonged lengths of stay, increased morbidity, and excess mortality.^3,4^ Increases in ED wait times also decrease patient satisfaction; patients are more likely to leave without being seen by a physician. While the consequences of crowding are well-documented, its causes are less well understood. A systematic review of over 100 studies found that only 14% examined the causes of crowding, while the majority focused on consequences and interventions.^5^

The input-throughput-output framework remains the dominant conceptual model for understanding ED crowding.^6^ Known drivers of throughput delays include high patient volume, inadequate staffing, prolonged boarding of admitted patients, and increasing patient complexity, factors that have been the focus of most operational improvement efforts to date. Environmental factors, including ambient temperature, have received little attention as potential determinants of ED throughput. Extreme heat is a well-documented driver of ED visit *volume*: numerous epidemiological studies consistently link high ambient temperatures to increased presentations across multiple diagnostic categories, including at the facility where the current analysis was conducted.^7,8,9^ The effect of extreme heat on events within the ED after patients arrive remains minimally characterized outside disaster-level heatwave situations.^10,11^ Under scenarios of continued climate change, dangerous heat events are projected to increase in both frequency and severity, potentially further straining ED operations at a time when the system is already under significant pressure.^12,13^

Characterizing the operational implications of heat for emergency care systems is an essential step toward building resilient healthcare systems. However, whether extreme heat disrupts operational performance within the ED, alters patient behavior, or shifts patterns of emergency service utilization beyond simply increasing the volume of ED arrivals remains unknown. Here, we examined the association between maximum daily heat index and ED throughput metrics, patient behavioral outcomes, and emergency service utilization among 485,000 warm-season visits to a tertiary care center ED over a 22-year period.

## METHODS

### Study Setting and Data Source

We analyzed ED visit data from a tertiary care center in New England, encompassing all Emergency Department patient encounters from 2002 to 2023, restricted to warm-season visits, defined as May 1 through September 30 of each calendar year. This study used de-identified data only and was deemed exempt from IRB Review (BIDMC IRB Protocol #2024P000981).

### Heat Exposure Data

We derived population-weighted daily maximum heat index values for Suffolk County, Massachusetts, from the ERA5-Land gridded dataset.^14^ (Supplemental Figure 1) Each ED visit was assigned the corresponding maximum daily heat index value for the date of arrival. Heat index incorporates both temperature and relative humidity and therefore better reflects the physiological burden of heat on human health compared to ambient temperature. The National Weather Service relies on heat index when issuing public heat advisories.^15^

### ED Visit Data and Throughput Metrics

We computed four emergency department operational time metrics using electronic timestamp data. For each ED visit in the warm-season dataset, we determined the total length of ED stay (LOS), door-to-doctor time, doctor-to-decision time, and ED boarding time (if applicable). LOS was defined as the time from the patient’s arrival at the ED to their physical departure. Door-to-doctor time was defined as the time from patient arrival to a physician’s first assignment to the patient. Doctor-to-decision time was defined as the time from physician assignment to the clinical decision regarding patient disposition. Boarding time was defined as the time from the disposition decision to physical departure from the ED for admitted patients. Given that ‘boarding’ applies specifically to admitted patients awaiting inpatient bed placement, rather than to all ED visits, we reviewed all disposition categories in the dataset to identify admitted patients and restricted boarding time calculations to this subset. We excluded operational time values that were negative or deemed implausible and likely attributable to data entry or documentation errors.

### Primary Analysis

We modeled each of the four throughput metrics as the outcome in separate generalized linear models using a Gamma distribution with a log link. The Gamma distribution was chosen to model the right-skewed, strictly positive distribution of operational time outcomes (Figure 1). Given that certain outcomes (e.g., door-to-doctor time for critically ill patients brought in by ambulance or helicopter and registered at bedside during initial physician evaluation) were zero, a small constant (0.000001 minutes) was added to all values across the four metrics to facilitate statistical analysis. The maximum daily heat index was treated as a continuous exposure variable. All models were adjusted for age, sex, day of week, month, and calendar year. Although age and sex do not confound the relationship between ambient heat and throughput, they were included as covariates to improve precision, given their known associations with ED operational outcomes. We did not adjust for daily visit volume or for variables likely to mediate the relationship between heat and throughput delays, such as ESI (Emergency Severity Index), imaging orders, and specialist consultation rates, as conditioning on mediators would have caused some degree of overadjustment bias. Calendar year was modeled as a categorical variable to account for long-term trends in typical weather, ED capacity, staffing, and workflows over the 22-year period. Results are expressed as the percent change in expected outcome time per °F increase in heat index. All analyses were conducted in R (Version 4.4.3, R Foundation for Statistical Computing) using RStudio.^16^

**Figure 1.**
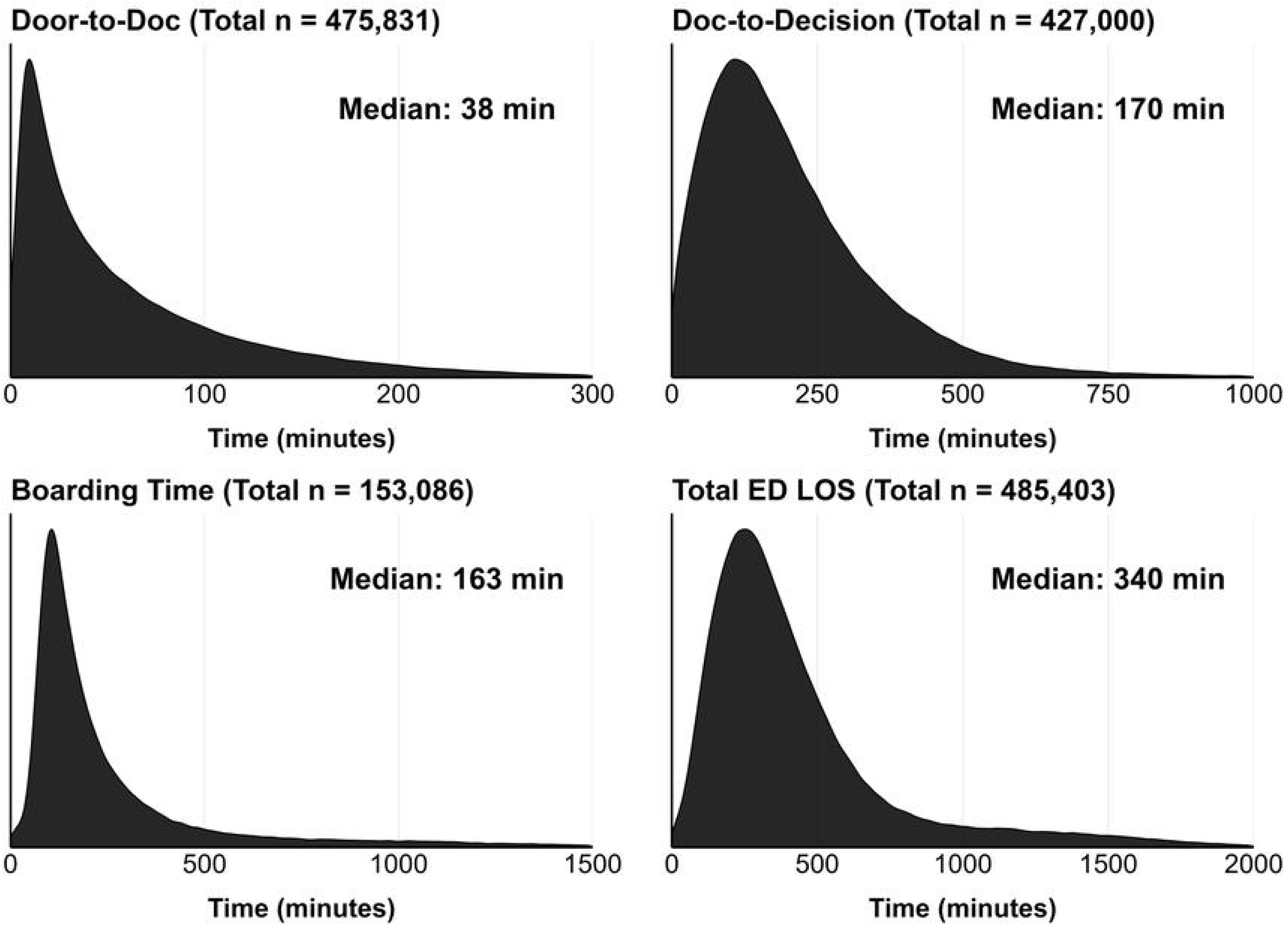
Distributions of the four ED operational time metrics (each metric exhibits a marked right skew). The labels in each panel show the median value.

### Secondary Analysis

To facilitate operational translation of our findings, we additionally dichotomized the maximum daily heat index at 90°F and 95°F (Supplemental Figure 1), corresponding to the National Weather Service ‘extreme caution’ classification and the Boston-area heat advisory threshold, respectively.^15,17,18^ Because such thresholds vary considerably by region and are not always grounded in local health outcome data, we use them here as operationally meaningful reference points rather than definitive markers of health risk.^19^ The results from binary models are expressed as a percent change in expected outcome time compared to days below each respective threshold.

We also examined two patient behavioral outcomes: left without being seen (LWBS) and left against medical advice (AMA). LWBS is defined as a patient’s departure from the ED prior to physician evaluation. AMA is defined as a patient’s departure after physician contact but before the recommended completion of care (referred to as ‘self-directed discharge’ in some health systems). Both outcomes were identified by reviewing all disposition categories in the electronic health record and manually coding visits. LWBS and AMA were modeled separately using logistic regression, with the daily maximum heat index dichotomized at 90°F and 95°F as the primary exposure. All models were adjusted for the same set of covariates as the primary analyses (age, sex, day of week, month, and calendar year). Results are expressed as odds ratios with 95% confidence intervals.

We additionally analyzed whether the likelihood of EMS arrival differed on extreme heat days. Logistic regression models were used separately for the 90°F and 95°F daily maximum heat index thresholds, adjusted for the same covariates as above (age, sex, day of week, month, and calendar year).

Door-to-doctor time was the only throughput metric with a statistically significant association with maximum daily heat index across all three models; we therefore conducted post hoc sensitivity analyses to assess the robustness of this finding. We independently refit the primary continuous heat index model with three additional covariates: same-day ED visit volume, prior-day ED visit volume, and ESI treated as a factor variable.^20^ These analyses evaluated whether the observed association was attenuated by adjustment for ED visit volume or patient acuity.

## RESULTS

The dataset included 485,403 warm-season (May through September) ED visits from 2002 to 2023. Patient demographic information for the overall dataset, and for the dichotomized dataset at the two thresholds, is displayed in Table 1. The distribution of maximum daily heat index values across the study period is shown in Supplemental Figure 1. During the study period, 35,787 visits occurred on days with a heat index of 90°F or higher, and 12,410 visits occurred on days with a heat index of 95°F or higher (Table 1). Not all metrics were available for every visit; door-to-doctor time was recorded for 475,831 visits, doctor-to-decision time for 427,000 visits, total ED LOS for 485,403 visits, and boarding time for 153,086 visits, the last reflecting the restriction to admitted patients only. (Supplemental Table 1) Median values and distributions of the four throughput metrics across the study period are presented in Figure 1.

**Table 1.** Demographic Characteristics of ED Visits.

| Characteristic | Overall<br>(n=485403) | HI<sub>max</sub> < 90°F<br>(n=449616) | HI<sub>max</sub> ≥ 90°F<br>(n=35787) | HI<sub>max</sub> < 95°F<br>(n=472993) | HI<sub>max</sub> ≥ 95°F<br>(n=12410) |
| --- | --- | --- | --- | --- | --- |
| **Age Category, n (%)** |  |  |  |  |  |
| <18 | 32 (0.0) | 29 (0.0) | 3 (0.0) | 29 (0.0) | 3 (0.0) |
| 18–44 | 190610 (39.3) | 176626 (39.3) | 13984 (39.1) | 185829 (39.3) | 4781 (38.5) |
| 45–64 | 152162 (31.3) | 140908 (31.3) | 11254 (31.4) | 148287 (31.4) | 3875 (31.2) |
| 65–74 | 61465 (12.7) | 56920 (12.7) | 4545 (12.7) | 59822 (12.6) | 1643 (13.2) |
| ≥75 | 81134 (16.7) | 75133 (16.7) | 6001 (16.8) | 79026 (16.7) | 2108 (17.0) |
| **Sex, n (%)** |  |  |  |  |  |
| Female | 262212 (54.0) | 243070 (54.1) | 19142 (53.5) | 255507 (54.0) | 6705 (54.0) |
| Male | 223191 (46.0) | 206546 (45.9) | 16645 (46.5) | 217486 (46.0) | 5705 (46.0) |
| **Race/Ethnicity, n (%)** |  |  |  |  |  |
| White | 293570 (60.5) | 272112 (60.5) | 21458 (60.0) | 286138 (60.5) | 7432 (59.9) |
| Black/African American | 103109 (21.2) | 95469 (21.2) | 7640 (21.3) | 100509 (21.2) | 2600 (21.0) |
| Hispanic/Latino | 37462 (7.7) | 34725 (7.7) | 2737 (7.6) | 36522 (7.7) | 940 (7.6) |
| Asian | 19288 (4.0) | 17840 (4.0) | 1448 (4.0) | 18776 (4.0) | 512 (4.1) |
| Other/Unknown | 31974 (6.6) | 29470 (6.6) | 2504 (7.0) | 31048 (6.6) | 926 (7.5) |
| **ESI Level, n (%)** |  |  |  |  |  |
| 1 (Immediate medical attention) | 24586 (5.1) | 22758 (5.1) | 1828 (5.1) | 23977 (5.1) | 609 (4.9) |
| 2 (Emergency) | 148506 (30.6) | 137386 (30.6) | 11120 (31.1) | 144460 (30.5) | 4046 (32.6) |
| 3 (Urgent) | 265087 (54.6) | 245751 (54.7) | 19336 (54.0) | 258422 (54.6) | 6665 (53.7) |
| 4 (Nonurgent) | 44342 (9.1) | 41048 (9.1) | 3294 (9.2) | 43305 (9.2) | 1037 (8.4) |
| 5 (Minor) | 2879 (0.6) | 2670 (0.6) | 209 (0.6) | 2826 (0.6) | 53 (0.4) |
| **EMS/Helicopter Arrival, n (%)** | 172777 (35.6) | 159699 (35.5) | 13078 (36.5) | 168050 (35.5) | 4727 (38.1) |
| **Medicare Insured, n (%)** | 145232 (29.9) | 134636 (29.9) | 10596 (29.6) | 141541 (29.9) | 3691 (29.7) |
| **Admitted, n (%)** | 180773 (37.2) | 167283 (37.2) | 13490 (37.7) | 176031 (37.2) | 4742 (38.2) |

Note: Age was categorized into five groups: younger than 18, 18–44, 45–64, 65–74, and 75 years or older. Race and ethnicity were self-reported and collapsed into five categories: White, Black or African American, Hispanic or Latino, Asian, and Other or Unknown. Insurance status was characterized as a binary variable indicating whether or not the patient had any Medicare coverage, identified by string matching on primary insurance fields. Arrival by EMS was defined as transport by ground ambulance or air medical helicopter, identified using mode-of-arrival codes. ESI level was treated as a five-level categorical variable.

In the primary analyses where heat index was treated as a continuous exposure, each 1°F increase in the maximum daily heat index was associated with a 0.41% increase in door-to-doctor time (95% CI: 0.38% – 0.44%) (Table 2).

**Table 2.** Association Between Maximum Daily Heat Index and ED Operational Metrics.

| Metrics on Mode of Arrival | Metric | Odds Ratio<br>≥90°F vs. <90°F<br>(95% CI) | Odds Ratio<br>≥95°F vs. <95°F<br>(95% CI) |  |
| --- | --- | --- | --- | --- |
|  | EMS Arrival | 1.05<br>(1.02 – 1.07) | 1.10<br>(1.06 – 1.15) |  |
| Throughput Metrics | Metric | % Change on Days ≥90°F vs. <90°F<br>(95% CI) | % Change on Days ≥95°F vs. <95°F<br>(95% CI) | % Change per 1°F Increase in Heat Index<br>(95% CI) |
|  | Door-to-Doctor Time | 3.05%<br>(1.89% – 4.21%) | 2.04%<br>(0.19% – 3.92%) | 0.41%<br>(0.38% – 0.44%) |
|  | Doctor-to-Decision Time | 1.35%<br>(0.38% – 2.32%) | 0.33%<br>(-1.37% – 2.06%) | 0.01%<br>(-0.02% – 0.03%) |
|  | Boarding Time | 1.53%<br>(-1.15% – 4.29%) | 3.15%<br>(-1.18% – 7.67%) | 0.05%<br>(-0.03% – 0.14%) |
|  | ED Length of Stay | 0.64%<br>(-0.46% – 1.75%) | 0.85%<br>(-0.93% – 2.66%) | 0.03%<br>(-0.01% – 0.06%) |
| Behavioral Metrics | Metric | Odds Ratio<br>≥90°F vs. <90°F<br>(95% CI) | Odds Ratio<br>≥95°F vs. <95°F<br>(95% CI) |  |
|  | LWBS<sup>a</sup> | 1.06<br>(0.98 – 1.15) | 0.96<br>(0.84 – 1.10) |  |
|  | AMA<sup>b</sup> | 1.11<br>(0.96 – 1.29) | 1.27<br>(1.02 – 1.58) |  |
| Secondary Analyses of Door-to-Doctor Time | Model | % Change per 1°F Increase in Heat Index<br>(95% CI) | Secondary analyses were conducted using the continuous heat index exposure model only |  |
|  | Adjusting for Prior Day Visit Volume | 0.39%<br>(0.36% – 0.42%) |  |  |
|  | Adjusting for Same-Day Visit Volume | 0.11%<br>(0.08% – 0.15%) |  |  |
|  | Adjusting for Emergency Severity Index | 0.39%<br>(0.36% – 0.42%) |  |  |

<sup>a</sup>LWBS = Left Without Being Seen (when a patient departs from the ED without being evaluated by a physician)
<sup>b</sup>AMA = Against Medical Advice (when a patient is not medically ready for discharge from the ED, but opts to leave; this is also known as self-directed-discharge in some health systems)

Visits occurring on days with a maximum heat index at or above 90°F were associated with a 3.05% increase in door-to-doctor time compared to cooler days (95% CI: 1.89% – 4.21%), with a smaller but statistically significant increase in doctor-to-decision time (1.35%; 95% CI: 0.38% – 2.32%) (Supplemental Figure 2). At the 95°F threshold, door-to-doctor time remained statistically significantly elevated (2.04%; 95% CI: 0.19% – 3.92%); doctor-to-decision time had a similar direction, but this trend was no longer statistically significant (0.33%; 95% CI: - 1.37% – 2.06%) (Supplemental Figure 3).

The association between maximum daily heat index and door-to-doctor time was minimally affected when adjusting for ESI (0.39% increase in door-to-doctor time per 1°F increase in heat index, 95% CI: 0.36% – 0.42%) and prior-day visit volume (0.39% per 1°F; 95% CI: 0.36% – 0.42%). (Table 2) Adjustment for same-day ED visit volume attenuated but did not eliminate the association, which decreased from 0.41% to 0.11% (95% CI: 0.08% – 0.15%) increase in door-to-doctor time per 1°F rise in heat index with this adjustment, suggesting that increased door-to-doctor time in hot weather is only partially explained by increases in ED arrival volume.

We additionally considered two patient behavioral outcomes: LWBS (n = 7896) and leaving AMA (n = 2393) (Table 2). At the 95°F heat index threshold, LWBS rates were not significantly different (OR 0.96; 95% CI: 0.84 – 1.10); however, patients were 27% more likely to leave against medical advice than on days below the threshold (OR 1.27; 95% CI: 1.02 – 1.58). Heat was associated with a significantly higher likelihood of EMS arrival at both thresholds. (Table 2) On days with a maximum heat index at or above 90°F, patients had 5% higher odds of arriving by ground ambulance or medical air transport compared to cooler days (OR 1.05; 95% CI: 1.02 – 1.07); this association strengthened at the 95°F threshold (OR 1.10; 95% CI: 1.06 – 1.15).

## DISCUSSION

We examined the association between daily maximum heat index and ED operational performance across seven outcomes, among nearly 500,000 warm-season visits to a large urban academic medical center over a 22-year period. Daily maximum heat index was associated with higher door-to-doctor time; this finding was robust across three modeling approaches, as well as sensitivity analyses. While associations with other throughput metrics were directionally consistent, they remained largely non-significant. The door-to-doctor finding was minimally affected by adjustment for ESI or prior-day visit volume but was substantially attenuated by adjustment for same-day visit volume, indicating that heat’s effect on door-to-doctor time is partially explained by concurrent increases in ED arrivals. The behavioral outcomes tell a complementary story: while LWBS rates were not significantly elevated on hot days, patients on days with a maximum heat index at or above 95°F had 27% higher odds of leaving AMA. Extreme heat was also associated with a shift in how patients arrived: the odds of a patient arriving by EMS were 5% higher on days at or above 90°F and 10% higher on days at or above 95°F compared to cooler days.

While modest at the individual level, the cumulative effect of increasing door-to-doctor time across a realistic temperature range is clinically and operationally meaningful. An increase in heat index from 70°F to 90°F corresponds to an approximately 8% longer door-to-doctor time, or roughly 5 additional minutes per patient. The *cumulative* impact across thousands of patient visits and repeated hot days may meaningfully strain health system resources over time.

Prior research has linked extreme heat to increased ED presentation volume.^7^ Our findings add new dimensions: heat also disrupts operational performance within the ED and alters both how patients arrive and how they behave once evaluated, thereby adding to the evidence base needed to develop systems-level solutions.^21^

These findings also underscore the importance of integrating environmental data into operational planning to strengthen climate resilience, and call for collaboration among public health leaders, local officials, and healthcare systems to proactively manage climate-related disruptions.^22,23^ Efforts to develop and implement clinical climate adaptation toolkits, hot weather alerts, and other patient and clinician-facing resources are ongoing.^24,25^ Affected communities and health systems must move beyond reactive responses and develop proactive strategies to prepare for and withstand the growing burden of heat.^21,26^

Heat’s operational impacts appear concentrated at the beginning of the patient visit, as reflected in arrival mode and door-to-doctor time. The lack of a statistically significant association with doctor-to-decision time suggests that once physician contact is made, the pace of clinical workup is not detectably slowed in this sample. This observation raises questions about whether downstream resource demands differ on hot days. This pattern suggests that interventions to mitigate the operational impact of heat should prioritize triage and the early stages of an ED visit.

The findings of this study also suggest a critical misalignment between public health warning systems and the operational needs of the health system. ED performance begins to degrade at a heat index of 90°F, five degrees below the threshold for a heat emergency in the City of Boston.^18,17^ Public health heat advisories serve a population-level protective goal; they are not designed as operational triggers for health systems. This study suggests that hospitals may need *facility-specific* heat thresholds, informed by their own operational data, to activate staffing and workflow protocols before a public advisory is declared; the binary models presented here convert a continuous dose-response relationship into actionable, threshold-based guidance. Operationally relevant thresholds will vary by institution and location, and will require periodic recalibration as ambient temperatures continue to rise and interventions are deployed in response.

The unit of analysis in this study was individual patient visits, and the associations reported in our results are modest due to this per-patient effect. However, their true significance at the systems level emerges when assessing the cumulative burden across thousands of visits. In this sample, there were 35,787 visits (on average 1,627 per year) on days with a heat index at or above 90°F. Applying this adjusted effect suggests an additional 45 cumulative *days* of door-to-doctor waiting time attributable to heat exposure over the study period; an approximation, but one that illustrates how a modest per-visit delay compounds into a substantial systems-level burden. The effects here represent a single data point in what may be a nationwide pattern of heat-related operational strain across healthcare systems. The burden of these operational disruptions will not be distributed equally.^27^ Older adults, patients with chronic conditions, and communities with limited access to primary care account for a disproportionately high share of ED visits.^28,29^ These populations significantly overlap with those that face greater risk during hot weather, making them uniquely vulnerable to the compounding effects of heat and ED operational strain.^30^

This study contributes empirical data across multiple nodes of the framework depicted in Figure 2, including pre-hospital EMS transport, ED operational metrics, and patient behavioral outcomes. However, much of the continuum, including inpatient operations and post-discharge recovery, remains poorly characterized. The analytic approach demonstrated here provides a replicable template for conducting such investigations across diverse health system settings.

**Figure 2.**
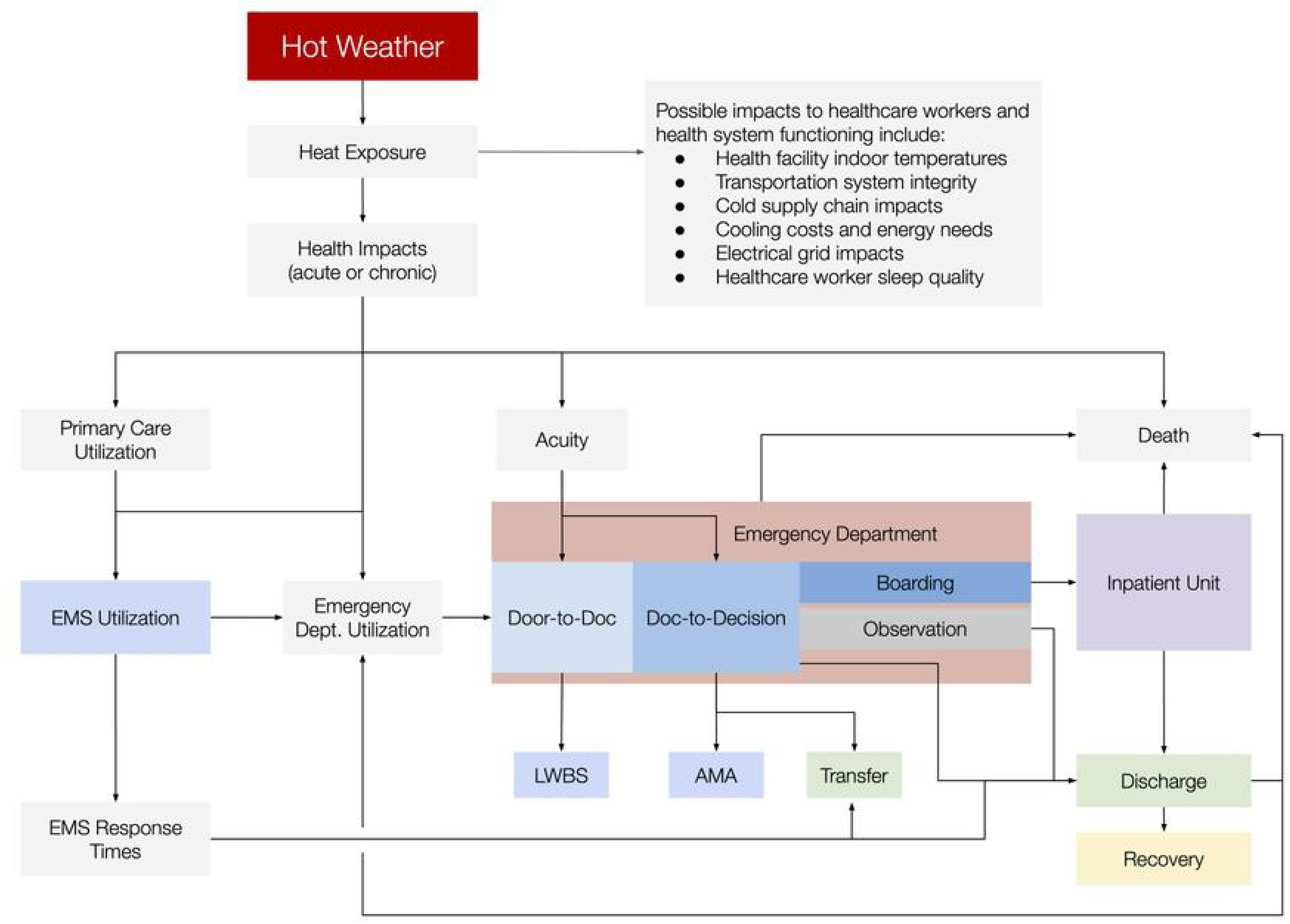
Conceptual framework for characterizing the impact of heat on healthcare systems functioning. The operational impact of heat on health system throughput and patient behavior studied here is only a component of the potential impact that heat may have on health system functioning. Metrics assessed in this article are shaded blue. Prior research has begun to characterize aspects of this framework, but most pathways through which heat may affect health system functioning remain uncharacterized; integrated analyses may provide additional insights. The ED represents a single node on a continuum spanning community-level heat exposure, emergency medical services delivery, acute and inpatient care, and patient recovery.

The specific numerical findings reported here may not be directly generalizable to other health systems, as the analysis is based on a single-center study conducted at a well-resourced academic medical center; impacts may be larger in less well-resourced environments. In addition, health systems in regions with more extreme or year-round heat exposure may experience operational impacts different from those observed here, due to differences in heat intensity, population acclimatization and vulnerability, or health system capacity and adaptation. However, the analytic approach presented here is generalizable and enables other healthcare systems to assess how region-specific climate variables may influence their operational performance. The temperature range in this analysis includes relatively few extreme heat days (above 95°F), approximately four days per year, limiting our statistical power to draw conclusions about the effects of extreme temperatures. Future work should examine the influence of additional types of adverse conditions (such as snow, pollution, precipitation, and wind) to better understand how changing weather patterns may affect ED operations. Additionally, the binary threshold approach, though useful from a health administrator’s decision-making perspective, is less sensitive than the continuous exposure model.

## Conclusion

This study offers both a replicable analytical approach and a conceptual framework that enable health systems to systematically characterize and respond to the operational consequences of environmental exposures. Integrating environmental data into hospital operations will be essential to safeguard patient access to timely care, particularly for those most vulnerable to climate-related disruptions. Health systems must take steps today to ensure that timely emergency care remains accessible as environmental hazards intensify, and the approach presented here can be adapted to inform these efforts. Doing so is what it means to build a health system in which all people can truly be well.

## Data Availability

The underlying data is protected individual patient information and cannot be made publicly available.

https://zenodo.org/records/21852053

## Acknowledgements

The authors would like to express gratitude to Dr. Murray Mittleman for his support in the early conceptualization of this work.

## Author Contributions (CRediT)

Conceptualization: NSB, GW, CD

Data curation: NSB, BS, NJ, CD

Methodology & Analysis: NSB, QA, GW, CD

Supervision: GW, CD

Visualization: NSB, GW, CD

Writing – original draft: NSB, GW, CD

Writing – review & editing: NSB, QA, BS, NJ, GW, CD

## Supporting Information

**Supplemental Table 1.** Annual Visits, Heat Exposure Days, and Throughput Time Summaries.

| Year | Total Visits | Days ≥ 90F HI | Days ≥ 95F HI | Visits ≥ 90F HI | Visits ≥ 95F HI | Mean LOS | Median LOS | Mean Door-to-Doc | Median Door-to-Doc | Mean Doc-to-Decision | Median Doc-to-Decision | Mean Boarding | Median Boarding |
| --- | --- | --- | --- | --- | --- | --- | --- | --- | --- | --- | --- | --- | --- |
| 2002 | 20302 | 20 | 7 | 2725 | 924 | 362 | 283 | 74 | 56 | 1731 | 116 | 304 | 227 |
| 2003 | 19673 | 5 | 2 | 641 | 255 | 407 | 311 | 70 | 50 | 207 | 131 | 331 | 267 |
| 2004 | 19092 | 4 | 0 | 508 | 0 | 431 | 329 | 60 | 39 | 238 | 147 | 299 | 61 |
| 2005 | 19635 | 13 | 4 | 1669 | 537 | 428 | 338 | 64 | 42 | 257 | 161 | 252 | 192 |
| 2006 | 20565 | 9 | 6 | 1236 | 833 | 430 | 330 | 63 | 42 | 206 | 161 | 227 | 140 |
| 2007 | 21724 | 12 | 2 | 1689 | 272 | 401 | 316 | 59 | 38 | 201 | 158 | 210 | 131 |
| 2008 | 22550 | 8 | 1 | 1144 | 167 | 363 | 292 | 54 | 35 | 187 | 156 | 150 | 101 |
| 2009 | 22369 | 7 | 1 | 1043 | 161 | 365 | 298 | 57 | 38 | 180 | 155 | 157 | 110 |
| 2010 | 24163 | 14 | 3 | 2256 | 504 | 391 | 314 | 59 | 38 | 182 | 157 | 194 | 128 |
| 2011 | 23903 | 6 | 4 | 971 | 622 | 388 | 308 | 57 | 37 | 183 | 157 | 176 | 117 |
| 2012 | 23447 | 13 | 3 | 2046 | 467 | 380 | 292 | 47 | 29 | 166 | 142 | 196 | 129 |
| 2013 | 23509 | 15 | 6 | 2264 | 923 | 397 | 302 | 51 | 33 | 172 | 144 | 217 | 144 |
| 2014 | 23644 | 8 | 1 | 1284 | 148 | 422 | 325 | 56 | 37 | 187 | 157 | 232 | 152 |
| 2015 | 23941 | 7 | 0 | 1153 | 0 | 469 | 357 | 68 | 45 | 197 | 167 | 291 | 182 |
| 2016 | 23965 | 13 | 6 | 2107 | 968 | 483 | 364 | 61 | 40 | 207 | 174 | 300 | 187 |
| 2017 | 23801 | 5 | 0 | 849 | 0 | 521 | 393 | 66 | 45 | 215 | 180 | 361 | 230 |
| 2018 | 23330 | 20 | 10 | 3107 | 1524 | 530 | 389 | 70 | 47 | 213 | 180 | 371 | 225 |
| 2019 | 22349 | 10 | 5 | 1468 | 715 | 564 | 405 | 76 | 46 | 218 | 183 | 426 | 253 |
| 2020 | 17116 | 15 | 4 | 1941 | 543 | 543 | 379 | 41 | 19 | 240 | 206 | 315 | 171 |
| 2021 | 21551 | 14 | 7 | 2154 | 1072 | 616 | 412 | 51 | 26 | 249 | 217 | 476 | 256 |
| 2022 | 22114 | 17 | 9 | 2456 | 1313 | 622 | 413 | 53 | 30 | 250 | 216 | 528 | 281 |
| 2023 | 22660 | 7 | 3 | 1076 | 462 | 670 | 457 | 74 | 44 | 269 | 227 | 559 | 314 |
| **Overall** | **485403** | **242** | **84** | **35787** | **12410** | **462** | **340** | **60** | **38** | **208** | **170** | **300** | **163** |

**Supplemental Figure 1.**
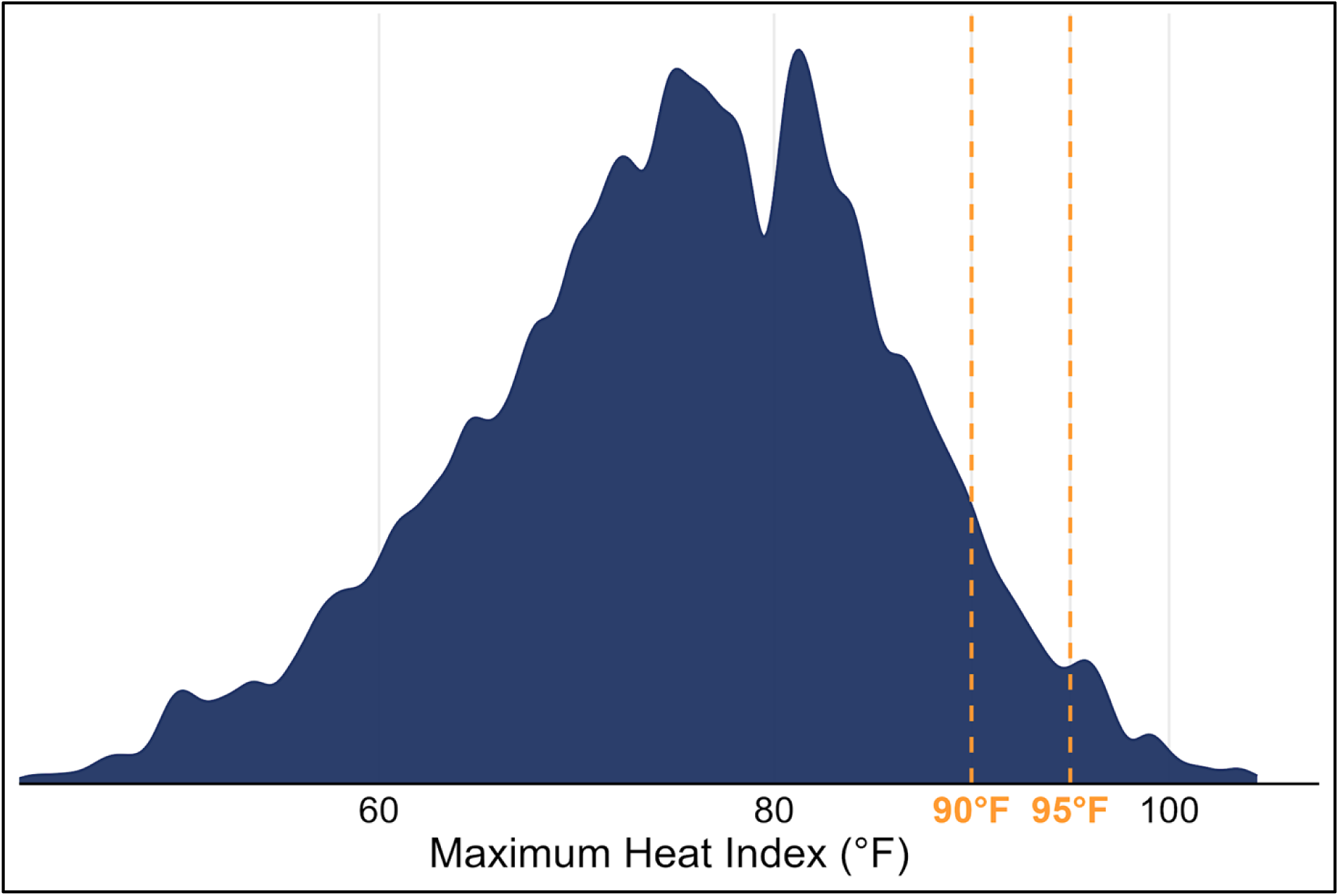
Density plot of maximum heat index during the warm season months (May–September), 2002–2023. The vertical dashed orange lines represent the 90°F and 95°F thresholds used in the analysis. During the study period, 35,787 visits occurred on days with a heat index of 90°F or higher, and 12,410 visits occurred on days with a heat index of 95°F or higher.

**Supplemental Figure 2.**
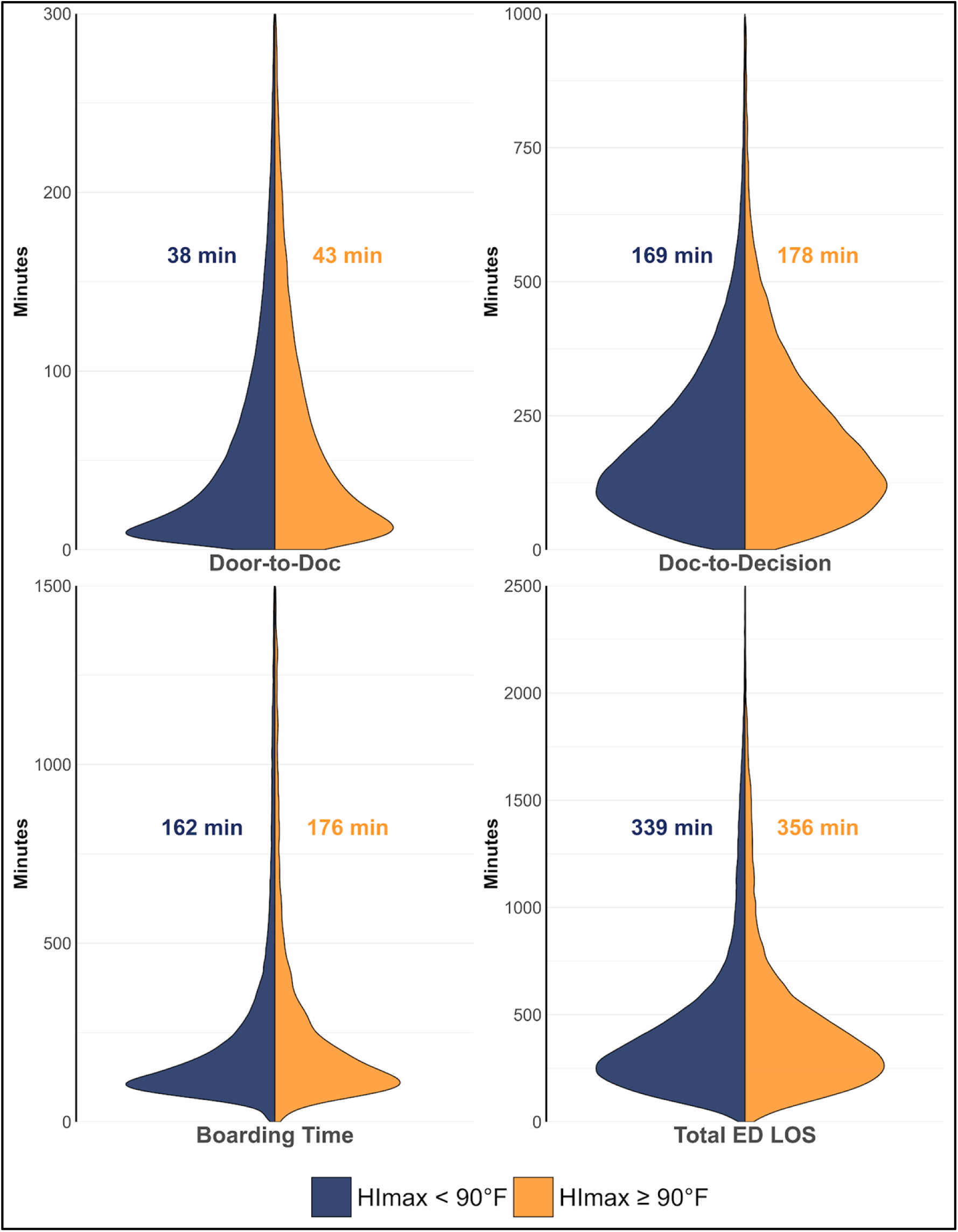
Split violin plots illustrating the distribution of the four ED throughput metrics of visits on days with a maximum heat index below 90°F versus at or above 90°F. The annotated values represent the median times calculated from the full dataset prior to y-axis truncation. Across all four metrics, the hotter day distributions show a consistent increase in throughput time relative to cooler days, reflecting a pattern of operational slowing on warmer days. Y-axes are truncated to highlight the primary distribution mass. Door-to-doctor time was the only throughput metric with a consistent, statistically significant association with heat index across all three models.

**Supplemental Figure 3.**
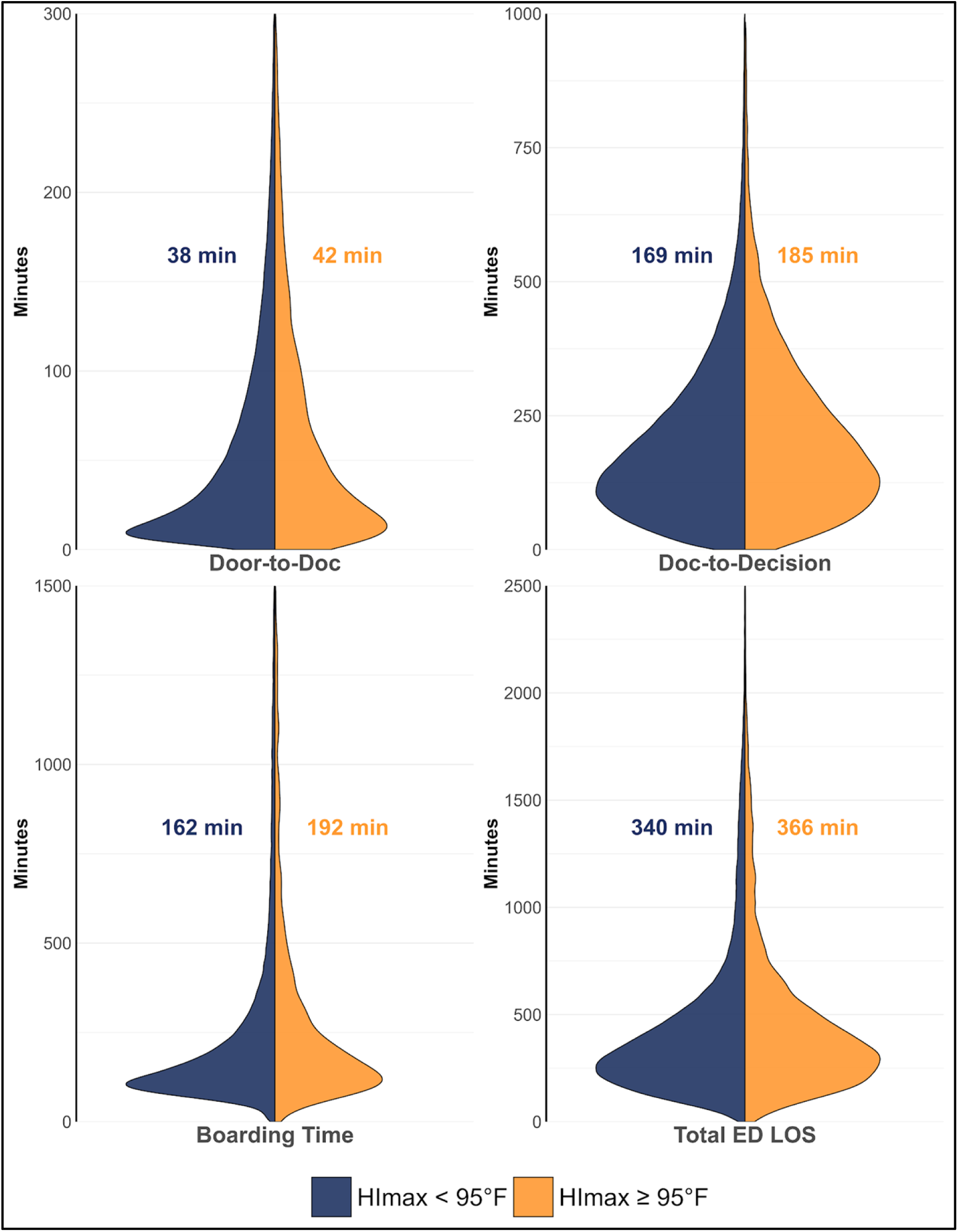
Split violin plots illustrating the distribution of the four ED throughput metrics of visits on days with a maximum heat index below 95°F versus at or above 95°F. The annotated values represent the median times calculated from the full dataset prior to y-axis truncation. Across all four metrics, the hotter-day distributions show a consistent increase in throughput time relative to cooler days, reflecting a pattern of operational slowing on extreme heat days. Y-axes are truncated to highlight the primary distribution mass. Door-to-doctor time was the only throughput metric with a consistent, statistically significant association with heat index across all three models.

